# A user-friendly, no-code, application for HIPAA-compliant automated analysis of tabular data at scale

**DOI:** 10.64898/2026.09.06.26362377

**Authors:** Paraic A. Kenny

## Abstract

**Background:** Clinical research often requires reviewing large volumes of unstructured electronic medical record (EMR) data, a time-consuming task demanding skilled personnel. Large language models (LLMs) like ChatGPT can efficiently analyze and summarize text, potentially accelerating chart review research. However, concerns about personal health information (PHI) leakage limit use of commercial chatbots. Secure, in-house LLMs within fire-walled environments can address these concerns.

**Aim:** To develop a user-friendly, scalable application leveraging an in-house ChatGPT 4o-mini on Emplify Health’s Azure cloud to automate processing of tabular clinical data while protecting PHI.

**Methods:** The application imports tabular data (Excel or text), guides users through prompt engineering, and automatically submits data row-by-row to the LLM. It retrieves and tabulates results as specified by users, enabling both automated extraction of clinical parameters, and data summarization/interpretation.

**Results/Conclusions:** Initially designed to identify cancer cases and extract related parameters from pathology reports, a fully generalized application was developed which has found utility in analyzing diverse data sources including cancer registries, cardiology CT reports, imaging narratives, and clinical notes. This tool has significantly accelerated research by reducing data retrieval time, allowing staff to focus on higher-value tasks like data analysis, while maintaining PHI protection.

## INTRODUCTION

Clinical research frequently involves the labor-intensive task of reviewing and extracting meaningful information from large volumes of unstructured electronic medical record (EMR) data (Murdoch and Detsky, 2013; Wang et al., 2018). This process often requires skilled personnel to manually curate and interpret complex clinical narratives, pathology reports, imaging descriptions, and other text-based records (Ford et al., 2016; Vassar and Holzmann, 2013). Such manual chart review is time-consuming, costly, and can delay research progress.

Recent advances in artificial intelligence, particularly large language models (LLMs) such as OpenAI’s ChatGPT, have demonstrated remarkable capabilities in natural language understanding and generation (Thirunavukarasu et al., 2023). These models can efficiently analyze, summarize, and extract relevant information from free-text data, offering the potential to dramatically accelerate clinical research workflows. However, widespread adoption of commercial LLM chatbots in healthcare settings is limited by concerns over patient privacy and the risk of personal health information (PHI) leakage (Marks and Haupt, 2023; Mesko and Topol, 2023), including the risk that future LLMs may be trained on the PHI of identifiable individuals (Carlini et al., 2021; Lehman et al., 2021).

To address these challenges, secure deployment of LLMs within fire-walled, HIPAA-compliant environments is essential. Such in-house solutions enable healthcare organizations to leverage AI-driven data analysis while maintaining strict control over sensitive patient data. While NLP applications have been developed (Savova et al., 2010; Soysal et al., 2018), to our knowledge user-friendly tools that allow non-technical clinical researchers to safely harness LLM capabilities for large-scale tabular data analysis remain scarce.

Here, we present a novel, no-code application designed to automate the processing of tabular clinical datasets using an in-house ChatGPT 4o-mini model hosted on Emplify Health’s Azure cloud. This tool guides users through prompt engineering, submits data row-by-row to the LLM, and retrieves structured outputs for downstream analysis. Initially developed for cancer case identification and pathology data extraction, the application has been generalized to support diverse clinical data sources, significantly reducing manual effort and accelerating research timelines while ensuring PHI protection.

## MATERIALS AND METHODS

### Application Development and Environment

We developed a user-friendly, no-code software application designed for automated, HIPAA-compliant analysis of tabular clinical data at scale. The application leverages an in-house deployed large language model (LLM), specifically the ChatGPT 4o-mini model, hosted on Emplify Health’s Azure cloud environment. This setup ensures secure processing within a fire-walled infrastructure to protect personal health information (PHI) while enabling scalable data analysis. A schema of the architecture is shown in Figure 1. Source code is available from https://github.com/paraickenny/AI_Tabular_Submit-Retrieve

**Figure 1.**
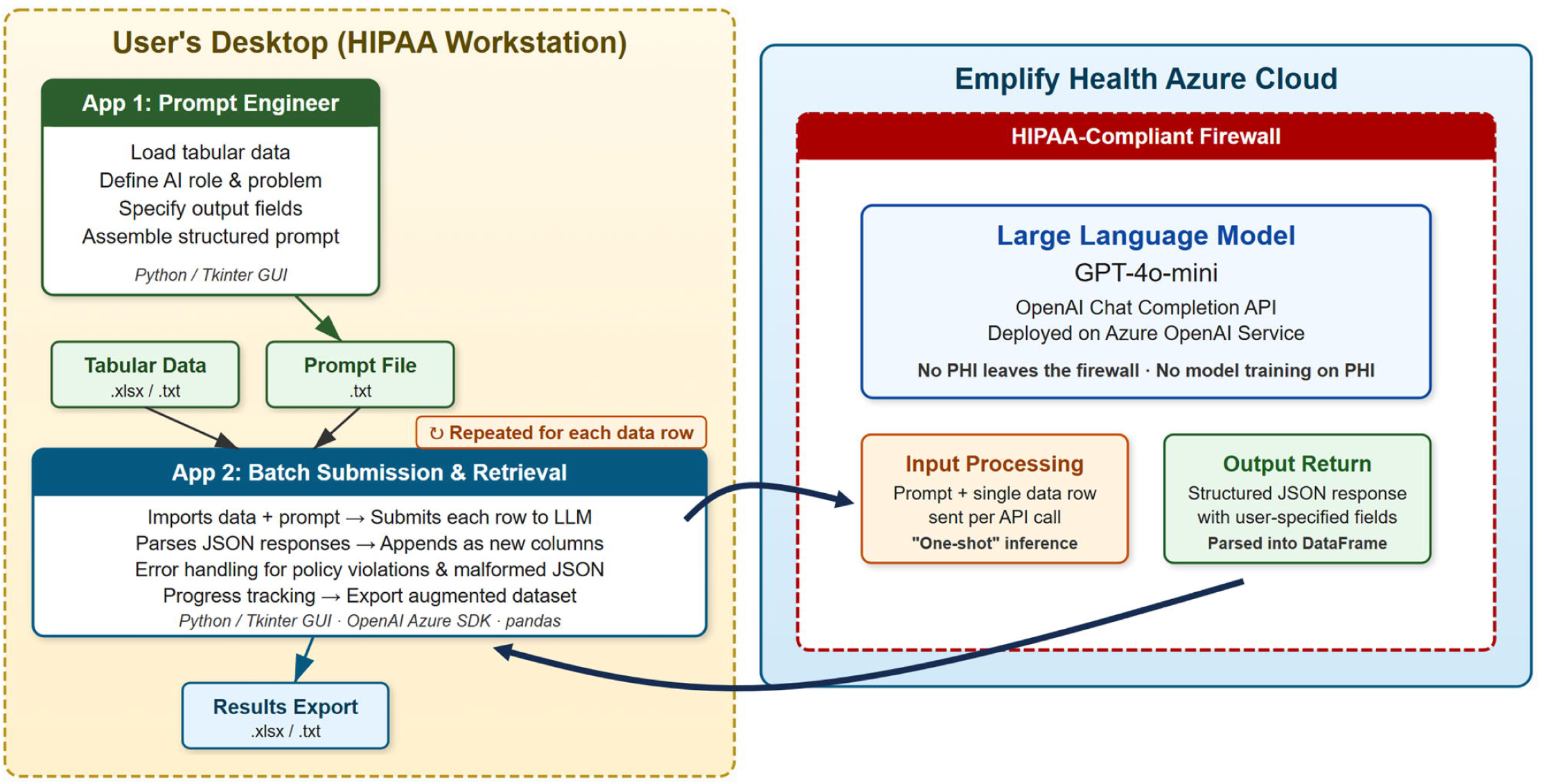
Application Architecture. Illustration of two-application workflow with data flow between user desktop and Azure cloud LLM within the secure, HIPAA-compliant, firewalled environment.

### Data Input and User Interface

The application supports importing tabular datasets in Excel (.xls, .xlsx) or tab-delimited text (.txt) formats. A graphical user interface (GUI) built with Python’s Tkinter library guides users through the process of prompt engineering. Users load their data, then provide detailed explanations of each data field to help the LLM understand the dataset context. The interface also collects user input on the role the LLM should assume (e.g., pathologist, cancer registrar) and a detailed description of the clinical problem to be addressed.

### Prompt Engineering

The application facilitates construction of a structured prompt that includes:

- The user-defined AI role
- Descriptions of each input data field
- The clinical problem statement
- Specification of desired output fields with detailed descriptions, including formatting instructions, if applicable

This prompt is assembled into a text format instructing the LLM to analyze each row of data and return results in JSON format without additional commentary. The prompt can be saved as a text file for use in batch processing.

### Batch Submission and Automated Analysis

A companion GUI application enables batch submission of tabular data to the Azure-hosted LLM. The application reads the user-prepared prompt and submits data row-by-row to the LLM endpoint using the OpenAI Azure SDK. Responses are parsed from JSON and appended as new columns to the original dataset. The interface displays progress and allows users to export the augmented dataset as Excel or tab-delimited text files.

### Error Handling and Quality Control

The batch submission tool includes error handling for common issues such as prompt content policy violations and JSON decoding errors. Errors are logged for review, and processing continues for remaining records to maximize throughput.

Implementation Details

- Programming Language: Python (v3.11)
- GUI Framework: Tkinter
- Data Handling: pandas (v2.23) library for reading, manipulating, and exporting tabular data
- LLM Access: OpenAI Azure SDK (v1.61.1) with environment variables for endpoint URL, API key, and model specification
- Model: ChatGPT 4o-mini deployed on Emplify Health’s Azure cloud
- Output Format: JSON for structured data extraction

To enable distribution across a potentially wide range of internal users, we separated the workflow into two applications – one for prompt engineering and one for data and prompt submission to the LLM. The latter application was password protected and had a remote kill switch which enabled it to be remotely shut down if any concerns about utilization or security arose. Distribution to end users was as zip files containing executables built with pyinstaller (v6.12.0).

### Use Cases and Validation

Initially developed to identify cancer cases and extract pathology report parameters, the application has been generalized and applied to diverse clinical datasets including cancer registries, cardiology CT reports, imaging narratives, and clinical notes. This tool has demonstrated significant acceleration of research workflows by automating data extraction while maintaining PHI security.

### Ethics and IRB Approval

The real-world use cases summarized in Table 1 draw on data from studies conducted at Gundersen Health System, primarily in the Gundersen Cancer Biobank. All studies were reviewed and approved by The Gundersen Clinic, Ltd. Human Subjects Committee/IRB under protocol numbers 2-24-06-001, 2-24-06-006, 2-22-02-003 and 2-25-08-005. All data were accessed and analyzed within the institution’s firewalled, HIPAA-compliant environment, and no personal health information left that environment at any point in the described workflows.

**Table 1.** A selection of real-world use cases from the Gundersen Cancer Biobank.

| Source Data | Approximate Number of Records | Problem | Notable challenges |
| --- | --- | --- | --- |
| Cancer registry records & biobank database entries | 14,000 | The Gundersen Cancer Biobank database contains rudimentary clinicopathologic data captured at the time of specimen accession, while the Gundersen Cancer Registry is a much richer data source, maintained longitudinally by a team of professional cancer registrars. Matching biobank records with the corresponding case records in the cancer registry would significantly improve the depth and quality of the annotation available in the biobank database and reduce the requirement for EMR chart review to retrieve data that may already have been captured within the organization. | Biobank data are "specimen-level" while cancer registry data are "case-level". The unifying ID was the patient's MRN. This resulted in a significant one-to-many cardinality issue where a patient may have had more than one cancer and/or multiple procedures such that a 1-to-1 crosswalk between the two datasets was not feasible. In addition, it was clear that terminology was used differently at several levels of both databases, e.g. "Lung" v "Bronchus", so fuzzy matching logic was required. |
| Pathology reports from endometrial cancer cases | 850 | Evaluate endometrial cancer cases to identify those with confirmed microsatellite instability testing and, of those, determine whether the testing protocol used met a study sponsor's requirements. | Evaluate pathology report, determine whether it relates to a biopsy or a resection, determine which cases had IHC or PCR assessment of microsatellite instability and, if IHC, determine whether the assessment was performed using two (PMS2/MSH6) or four (PMS2/MSH6/MLH1/MSH2) antibodies. If MSI tested, report result. If not tested, report "Not Tested". |
| Bulk pathology reports | 3000 | Evaluate pathology reports and identify cancer cases | Determine whether or not a report corresponds to a cancer specimen and, if so, extract multiple parameters of tabular data (e.g. tissue of origin, metastatic site (if any), IHC markers tested, tumor grade, etc.) |
| Provider visit notes | 1000 | Review free text notes to determine if doctor and patient had a discussion of drug toxicities | Recognition of drug names and NLP interpretation of whether the physician recorded a discussion of associated toxicities. |
| Pathology reports | 100 | Remove PHI for HIPAA-compliant data transfer to sponsor | Identify names of patients, providers, dates, MRNs, specimen IDs etc. and replace with [REDACTED]. |
| Pathology reports | 600 | Identify the optimal tumor-containing block for whole-slide imaging | Evaluate text of breast cancer pathology reports, including gross description, microscopic results, and any associated IHC results. We specified that the block chosen by the pathologist for ER/PR/HER2 testing was likely optimal for imaging. |
| MRI imaging notes | 100 | Analyze and deidentify 100 rectal MRI imaging notes. | Determine which elements correspond to PHI and date-shift all mentioned dates by a constant to preserve relative timing of events while protecting patient privacy. |
| Cancer registry pathology report summaries | 300 | Determine if a patient was treated with neoadjuvant chemotherapy and, if so, capture any recorded response data (e.g. RCB score). | Neoadjuvant therapy was often not explicitly described as such, and needed to be inferred by comparing treatment initiation date and surgery date. |

## RESULTS

### Summary of workflow with a synthetic use case

To evaluate performance of the application and prepare an example for user training, we requested ChatGPT generate a synthetic dataset of 100 patients, each with a name, a date of birth, a list of three medical problems as would be found in a patient’s electronic medical record, and the patient’s favorite animal (Fig 2Ai). Upon loading the test data set into the tool, the tool detects the column headers and creates a set of text boxes to enable the user to provide any clarifying information (Fig 2Aii). In this case, most of the fields are self-explanatory, but the user clarifies that the “favorite animal” should be interpreted as “Pet in the Home”.

**Figure 2.**
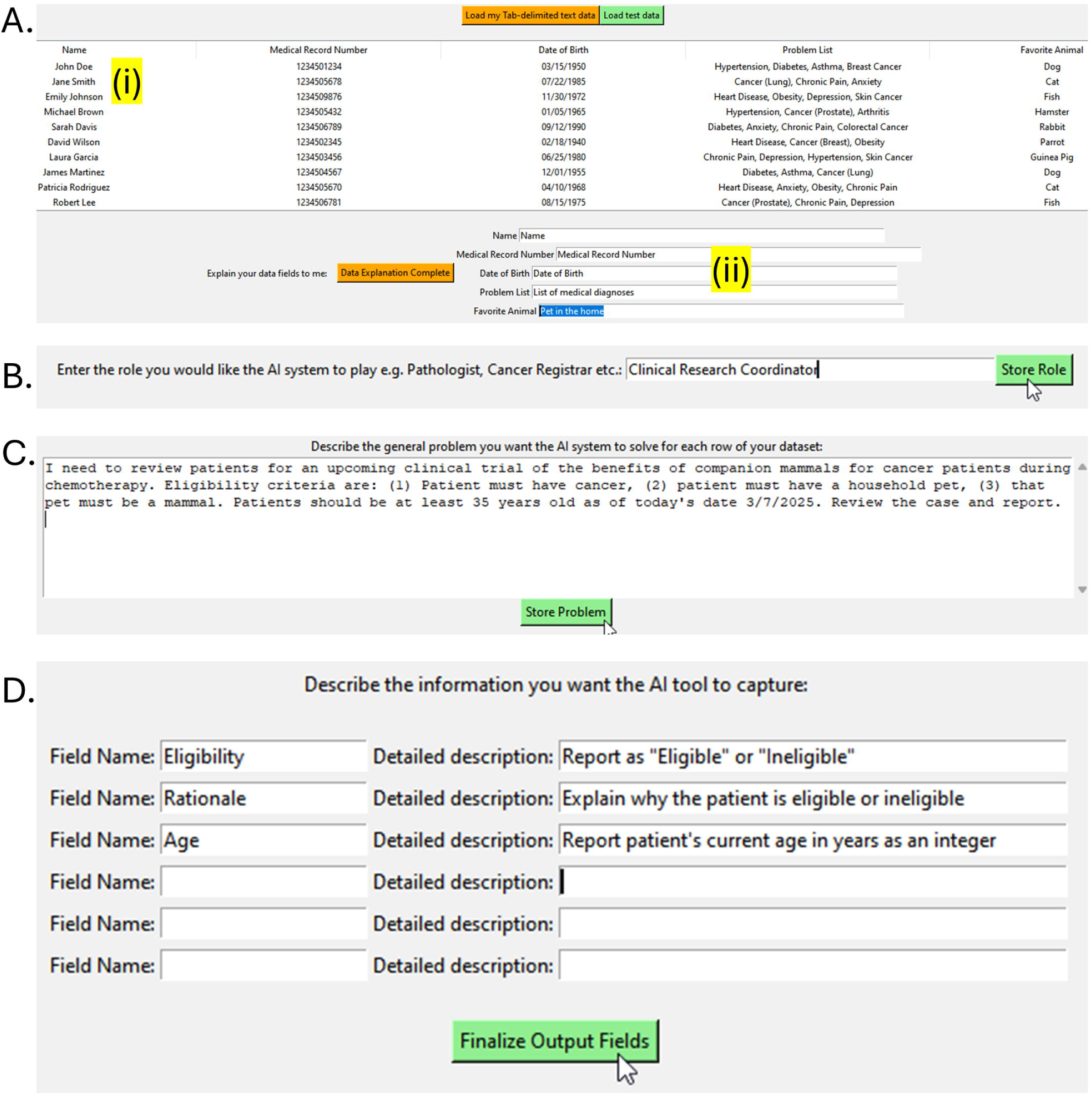
Illustration of prompt engineering application using a synthetic dataset. A. Synthetic dataset is loaded (i) and the user can enter clarifications for each auto-extracted column header (ii). B. Specification of the role that the LLM should play. C. The user describes parameters of the problem and any constraints. D. The user specifies what they want the LLM to determine for each row of data, and any additional constraints or formatting required in the output data.

The user then specifies the role that the LLM should play, in this case a Clinical Research Coordinator (Fig 2B), and then enters the problem that the LLM should solve for each row of the dataset (Fig 2C). For our demonstration problem, we specified that we were seeking to enroll cancer patients in a clinical trial who had a mammalian household pet and who were over 35 years old. Successful completion of this problem for each row of data would demonstrate reasoning and calculating abilities.

Finally, the user specifies the information that the LLM should capture by specifying fields for the output data and providing an explanation to the LLM of what it should report and any desired formatting. For example, we requested patient’s current age as an integer, to avoid the need to later harmonize responses like “42” and “forty two” prior to any subsequent analyses. In many use cases, we found it helpful to add a field for “Rationale” to better understand decisions made by the LLM so that prompting could be refined if the output did not meet our QC metrics.

When the user clicks “Finalize Output Fields”, the application assembles the input data into an engineered prompt (Fig 3) in which user-entered text is shown in black and the additional clarifying instructions, in red, are added by the tool. These supplemental cues provide additional guidance about returning the data in a format that the tool can easily parse (JSON) and requesting that no extraneous text or commentary should be returned. The text in the specific format “Output_Fields will be [item1, item2, item3, etc.]” encodes the information needed for the batch submission tool to create the necessary columns in the dataframe to store the returned information.

**Figure 3.**
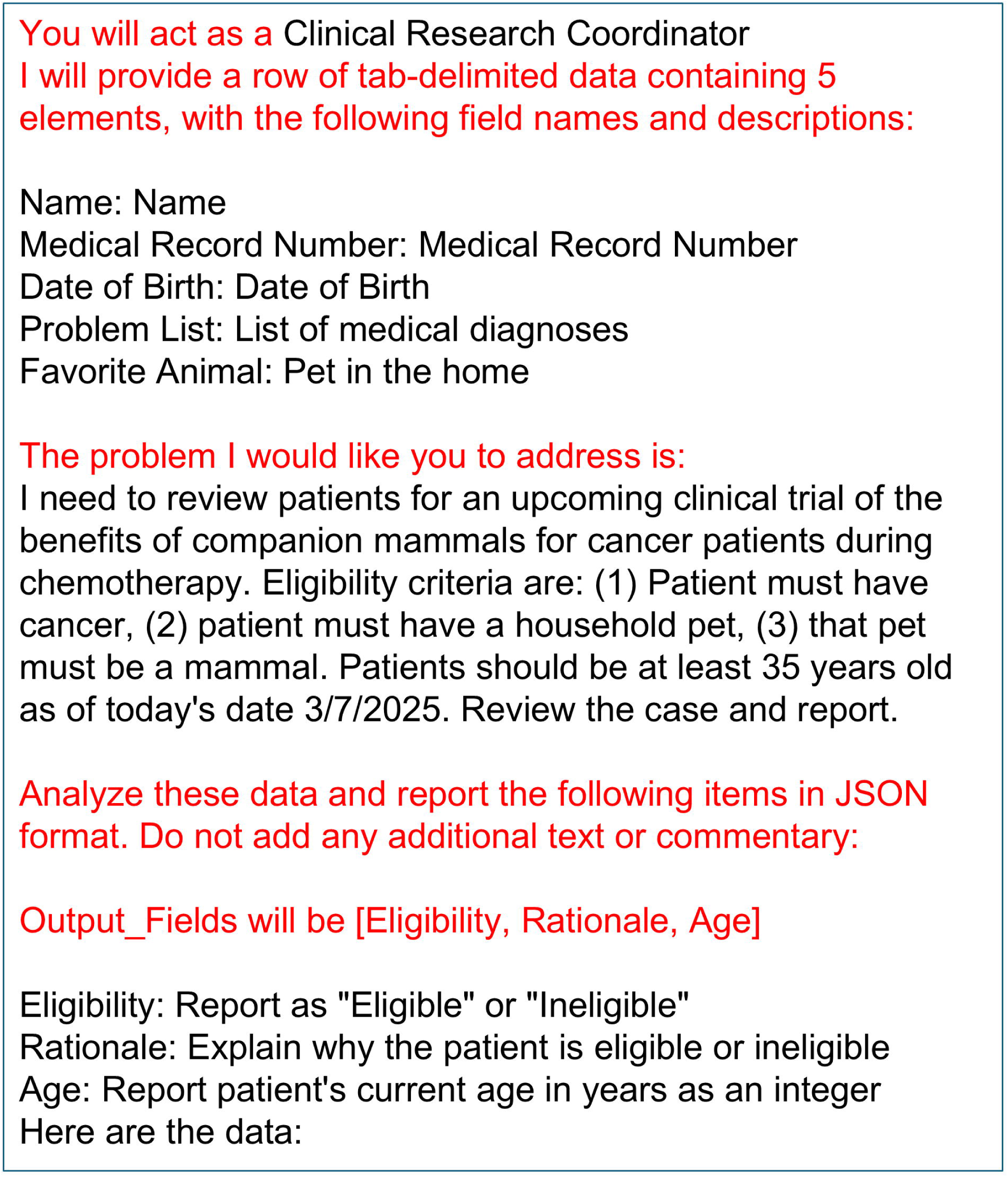
Prompt assembly by the application. User-provided text is in black. Additional instructions supplied by the prompt engineering tool are highlighted in red.

A second application is used to facilitate row-by-row submission of the engineered prompt and data to the LLM and to capture and parse the returned responses. (Fig 4). The original tab-delimited text data file is imported, along with the saved engineered prompt. The “Output_Fields” information encoded in the prompt specifies the column headers to add to the dataframe, and the process of data submission can be initiated with a click. The output for the first 10 rows of the data is shown in Figure 4. The results can then be exported in tab-delimited text or Excel format to enable quality control checks and subsequent data analysis or processing.

**Figure 4.**
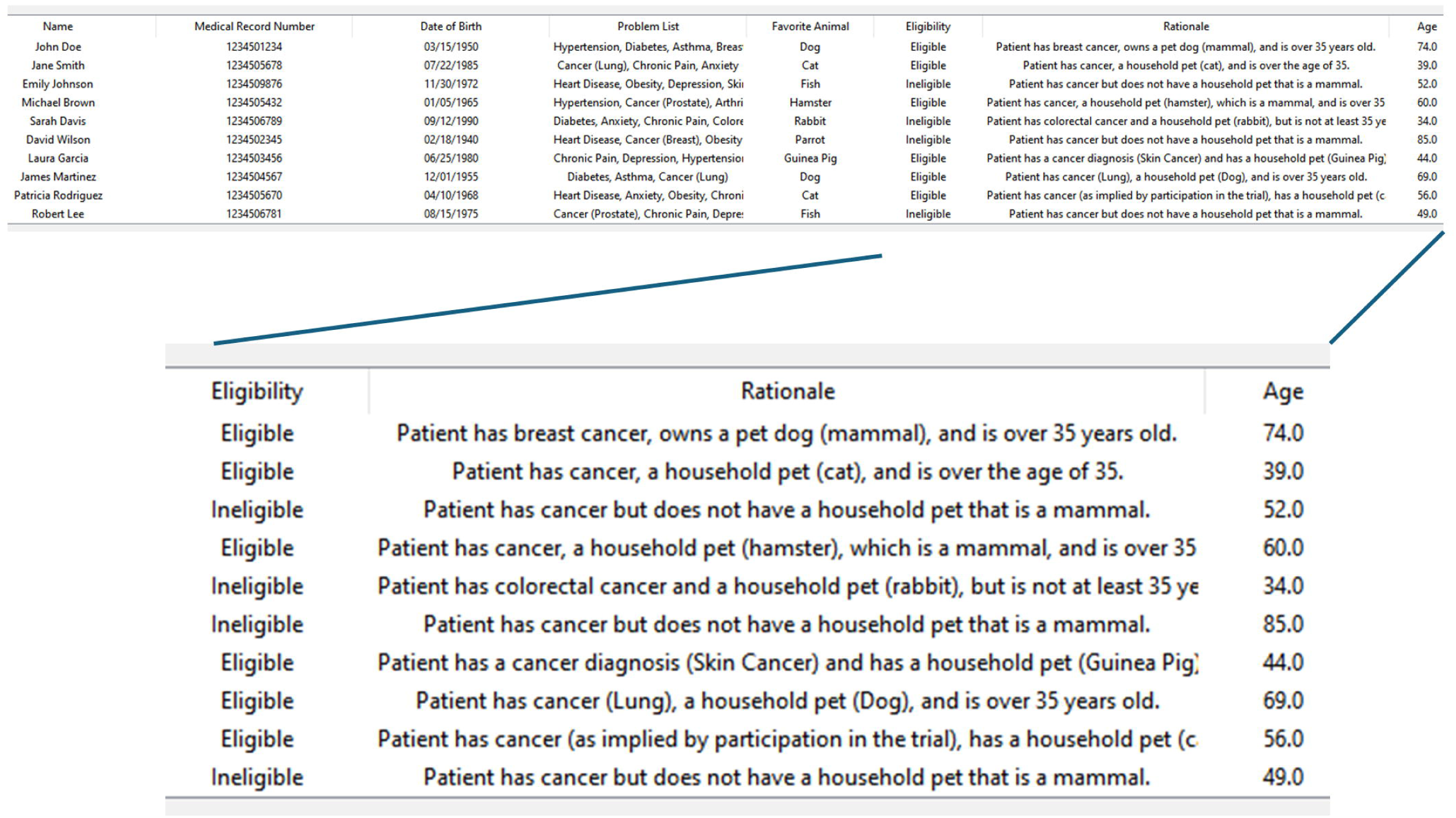
Screenshot of batch submission application. The prompt is submitted with each row of data, and the returned JSON response is parsed and added to the table. Upon completion, data can be exported in Excel or .txt format.

A summary of real-world examples of analyses performed is shown in Table 1. In all cases, the input format is tabular text data, so some simple pre-processing (e.g. copy/pasting or otherwise processing text files into a table) was required in some cases.

## DISCUSSION

We successfully built and deployed an application in our research division that uses an in-house cloud-based ChatGPT implementation for the automated processing of clinical text data at scale. Although many of the initial use cases were cancer-focused, other members of the research team used it for analysis of text notes from a wider variety of clinical settings, demonstrating broad applicability. The application was robust to large volumes of input data, and enabled the rapid acceleration of some projects which would have required days-to-weeks of human review, and also the performance of other projects that would not have been economically justifiable if performed at human pace and labor cost.

The first iteration of this approach was a command-line implementation specifically hard-coded to review pathology reports to abstract a specific and limited subset of named parameters. This required cloud access via the Databricks platform and the ability to sequentially execute code in Python notebooks. Although functional, the ability of typical users to adopt this approach successfully was considered to be low and widespread assignment of Databricks accounts for this purpose would have been challenging. Nevertheless, demonstration of the utility of this approach led to several requests from colleagues to build an application to address additional custom use cases. Rather than constantly build new applications, we instead focused on the entirely generalized and user-driven solution described herein. Providing a familiar graphical user interface was a key to ease-of-use (Davis, 1989), and we found that non-specialist users easily self-trained using a step-by-step PowerPoint guide provided. This underlines the importance of making technical advances accessible to non-specialist users.

Deployment of any new AI tool creates governance risks that need to be understood and mitigated, particularly in healthcare settings (Reddy, 2024). While an ever-increasing array of custom tools are being commercially developed, finding the institutional bandwidth to comprehensively review risks and engage with contracting represents a substantial headwind. For economic reasons, institutions will prioritize applications that are likely to be high-impact use cases for a broad user base, and that will hopefully either reduce cost or ease workflow bottlenecks. A network of users with idiosyncratic and rapidly evolving analytical needs, such as a group of clinical researchers, is unlikely to be well-served by existing models. A key advantage of our approach is that it harnessed an already well-vetted LLM implementation, and enabled our non-AI specialist user base to build and scale their own applications, using their own expertise and with their own research data. Reviewing the selected set of use cases in Table 1, all saved substantial time for individuals but none would rise to the level of becoming an institution-level implementation priority. Nevertheless, considering the time savings in aggregate, it is clear that the implementation of this approach saved substantial FTE across our division. In this way, the application led to cost savings, democratization of AI access and increased staff AI literacy and adoption.

A key failure point noted early in testing was errors returned by the LLM when confronted with phrases that violated one or more of the content filters. For example, the phrase “biopsy of necrotic rectal mass” was found in numerous pathology reports and triggered the “violence filter”. Some additional code was added for error handling for these and other situations, enabling stable completion of larger projects and logging of specific errors for user review at the completion of the batched analysis process.

One key limitation was that accuracy was heavily influenced by the user’s ability to clearly and precisely specify that they needed the LLM to abstract, interpret or summarize. Although no coding was required to use the application, a “coding mentality” was helpful. For most use cases, we suggested that users request a “rationale” as one of the output fields. This provided insight to the user about why the LLM made particular choices and, when those conflicted with user expectations, it enabled the user to revise the prompt to more clearly and unambiguously frame the problem to be addressed. Coupling this with a test set of 20-50 manually reviewed cases for each project (Ford et al., 2016) provided confidence in model performance prior to deployment across entire data sets.

Future enhancements may include AI-based feedback and guidance to users on prompt design and optimization, which currently requires user assessment of performance against calibrated data and adjustment of the prompt text, if needed. The current application also relies on provision of case data by the user to the LLM. A future iteration might include an agentic approach whereby an LLM trained on the structure of the EMR could directly retrieve and integrate discrete sources of data from various data silos, including clinical notes, pathology reports, lab results and imaging, prior to execution of a requested analysis of the assembled data. This would address a substantial bottleneck in clinical research. Deployment of such an approach over a standard EMR would present significant governance risks, but its use on a shadow, deidentified EMR could have substantial value.

In summary, we built and deployed a user-friendly application enabling HIPAA-compliant batched AI analysis of tabular data at scale which was widely adopted by a base of users without specialist computational skills. This resulted in significant acceleration of several clinical research projects at our institution.

## Data Availability

All source code is available online at https://github.com/paraickenny/AI_Tabular_Submit-Retrieve

## ACKNOWLEDGEMENTS

This work was supported by the Gundersen Medical Foundation. PK holds the Dr. Jon & Betty Kabara Endowed Chair in Precision Oncology. We thank Andrew Borgert and Krister Mattson for input and suggestions for this study.

## Notes

### Competing Interest Statement

The authors have declared no competing interest.

### Author Declarations

The Human Subjects Committee/Institutional Review Board of The Gundersen Clinic, Ltd. gave ethical approval for this work.

